# Cambodian *Plasmodium vivax* parasites with reduced hemoglobin digestion display delayed clearance upon artesunate treatment

**DOI:** 10.1101/2025.03.09.25323469

**Authors:** Kieran Tebben, Virak Eng, Dynang Seng, Baura Tat, Lionel Brice Feufack Donfack, Agnes Orban, Rominea Yeat, Jeremy Salvador, Sitha Sin, Katie Ko, Nimol Khim, Claude Flamand, Cecile Sommen, Dysoley Lek, David Serre, Jean Popovici

## Abstract

Artemisinin-based combination therapies are the frontline drugs for the treatment of malaria infections but, for *Plasmodium falciparum,* the efficacy of artemisinin is threatened by the spread of resistance*. P. vivax* is the second most common cause of human malaria but we have little information on its susceptibility to artemisinin due to the lack of *in vitro* cultures. Here, we analyze 161 *P. vivax* infections from Cambodian patients treated with 2 mg/kg/day of artesunate for seven days. All infections were successfully cleared by day 3. However, one third of the infections displayed a slow clearance after treatment, with nine infections (5.7%) with a parasite clearance time greater than 5 hours, meeting the WHO definition of artemisinin resistance. We observed no significant association between slow clearance and either patient- or infection characteristics (including stage composition). We used RNA-seq to characterize the gene expression of parasites from 15 fast- and 16 slow-clearing infections at baseline and 1, 2 and 4 hours after treatment. While fast-clearing parasites showed significant changes in gene expression immediately upon treatment, slow-clearing parasites displayed a significantly delayed gene expression response, with a downregulation of many genes associated with hemoglobin endocytosis and digestion. Overall, our results indicate that some Cambodian *P. vivax* parasites clear slowly after artesunate treatment, possibly due to a downregulation of hemoglobin metabolism that may reduce the efficiency of the artesunate.

**Research in context:** *Evidence before this study:* The WHO treatment guidelines recommend artemisinin-combination therapy (ACT) for treatment of blood-stage infections caused by *Plasmodium vivax* in all areas (with chloroquine recommended only in areas where *P. vivax* are still chloroquine-sensitive). In *P. falciparum*, partial resistance to artemisinin derivatives is defined *in vivo* as either detected parasitemia on day 3 post treatment or as a half-life of the parasite clearance slope of ≥ 5 hours. We searched Pubmed for studies containing the terms "vivax" AND "clearance" AND ("artesunate" OR "dihydroartemisinin" OR "artemether" OR "artemisinin") published between 1990 and February 2025, with no language restrictions. Our search retrieved 102 studies for which title and abstracts were screened to identify 21 studies reporting outcomes of *P. vivax* treatment with an artemisinin derivative. While all these studies concluded that artemisinin derivatives provided rapid clearance of *P. vivax* parasites, two studies reported a low frequency of day 3 positivity following artesunate-amodiaquine treatment (2.6% in Brazil) or dihydroartemisinin-piperaquine (0.6% in Indonesia). No study reported clearance slope half-life ≥ 5 hours.

*Added value of this study:* This study used a cohort of Cambodian patients infected by *P. vivax* to rigorously examine the efficacy of artesunate monotherapy at clearing blood stage infections. Our study showed significant variations in clearance rates among infections, with 5.7% of the infections with a clearance slope half-life ≥ 5 hours, meeting the criteria for artemisinin partial resistance used for *P. falciparum*. Variations in clearance rate upon artesunate treatment were not associated with patient or infection characteristics. Gene expression analyses revealed that the slow-clearing parasites down-regulated upon treatment many genes involved in hemoglobin endocytosis and digestion, possibly resulting in a lesser activation of artesunate.

*Implications of all the available evidence:* Our results confirm that 2 mg/kg of artesunate per day for seven days is effective at clearing *P. vivax* blood stage infections. However, a subset of the *P. vivax* parasites displayed a slow clearance following artesunate treatment meeting artemisinin partial resistance definition in *P. falciparum*. Gene expression analyses suggest that metabolic variations may underlie slow clearance. Increased monitoring of treatment efficacy and drug resistance in *P. vivax* is therefore recommended.

## Introduction

Artemisinin-based combination therapies (ACTs) are the recommended first-line treatment for uncomplicated malaria in all endemic areas, but the efficacy of the treatment is challenged by the emergence and spread of drug resistance. Delayed clearance of *P. falciparum* parasites after treatment with artemisinin was first reported in the 2000s in Western Cambodia [1, 2], before rapidly spreading throughout the Greater Mekong region [3]. Thanks to extensive control measures, the spread of these initial resistance alleles seems to have been successfully contained to Southeast Asia. However, independent emergence of artemisinin resistance has since been reported throughout the world, including in Africa where it could jeopardize the progress made towards malaria elimination [4–6]. Resistance to artemisinin in *P. falciparum* is more accurately defined as a delayed clearance upon treatment than a true resistance enabling the parasites to survive and grow in the presence of the drug. However, given the short half-life of artesunate in patients (less than 2 hours), delayed clearance is sufficient for parasites to outlast the drug and persist after treatment. Artesunate is therefore recommended to be used in combination therapy, with a long-lasting partner drug (e.g., with mefloquine, lumefantrine or piperaquine). Unfortunately, reports of resistance to artemisinin were rapidly followed by reports of resistance to the partner drugs (e.g., [7–9]), suggesting that the acquisition of delayed clearance to artesunate might facilitate the emergence of resistance to the partner drugs.

The mode of action of artesunate on *Plasmodium* parasites remains incompletely understood but it requires the activation of the drug by heme, generated by the digestion of hemoglobin by the parasite, and leads to widespread alkylation of proteins and generation of radical oxygen species and DNA damage [10, 11]. The mechanisms underlying artesunate resistance in *P. falciparum* are complex and multifaceted but likely involve decreased hemoglobin uptake and degradation, proteotoxic/unfolded protein stress response as well as oxidative stress response [12–15].

While *P. falciparum* is the main cause of malaria mortality and is responsible for most infections worldwide, *Plasmodium vivax* caused more than 69 million cases of malaria in 2022 and is a major public health concern given its resilience to elimination efforts [16]. ACTs are now increasingly used against uncomplicated vivax malaria (with dihydroartemisinin-piperaquine used in Cambodia after 2013 and artesunate-mefloquine since 2018). Antimalarial drug resistance in *P. vivax* seems less widespread than in *P. falciparum* (and more difficult to rigorously demonstrate due to the lack of a robust *in vitro* system and the confounding effect of relapses in patient studies, see e.g., [17]). Indeed, most cases of drug resistance in *P. vivax* have been reported much later than their descriptions in *P. falciparum* and often decades after the introduction of the drug [18, 19]. Here, we describe the analyses of 161 *P. vivax* mono-infections from 87 Cambodian patients treated with 2 mg/kg/day of artesunate for seven days and examine variations in parasite clearance times. We also use whole genome sequencing and gene expression analyses to evaluate the response of the parasites to artesunate treatment and to characterize the genetic and molecular factors influencing clearance time in *P. vivax*.

## Materials and Methods

### Study design and participants

All samples were collected from participants enrolled in a open-label clinical trial (NCT04706130) designed to evaluate the efficacy of two regimens of primaquine (0.25 or 0.5 mg/kg/day for 14 days) by comparing them to patients who did not receive primaquine.

Briefly, eligible individuals seeking treatment for malaria at one community or health center in the Kampong Speu province (Western Cambodia) and diagnosed by rapid diagnostic test with *Plasmodium vivax* infection were offered to participate in the study. All participants had uncomplicated vivax malaria (confirmed by qPCR), were aged 15 years or older, and had hemoglobin concentration of 8 g/dl or more. Pregnant and breastfeeding women were excluded from this study. All enrolled patients or their guardians provided written informed consent, and assent was obtained for all patients aged 15-18 years old. Ethical clearance was obtained by the National Ethics Committee for Health Research of Cambodia (158-NECHR, 06/29/2020) and the University of Maryland IRB (HP-00091095), and the study was overseen by the NIH Division of Microbiology and Infectious Diseases (Protocol 20-0010).

### Antimalarial drug treatment

All participants received 2 mg/kg/day of artesunate (RT Nate, manufactured by Bonn Schtering Bio Sciences, India) for seven days, starting on the day of enrollment. Randomization to one of the two primaquine regimen, or to the comparator arm without primaquine, was performed once G6PD activity was determined by spectrophotometry (by day 7). Primaquine administration, if any, was started after the end of artesunate therapy (on day 7).

### Procedures

Upon enrollment, a clinical examination was performed and a medical history was taken. Artesunate treatment (50 mg pre-scored tablets) was administered under direct supervision and adjusted to body weight. A venous blood sample was collected before the first dose of artesunate (0h) and capillary blood samples were collected by finger prick in EDTA microtainer tubes 1h, 2h, 4h, 8h and 16h after the first dose of artesunate, and then at 24h intervals during the seven days of artesunate therapy.

All participants were relocated to the town of Aoral (Kampong Speu province) for the entire follow-up duration (90 days) to ensure they were not reinfected. A capillary blood sample was collected every second day throughout the follow-up period. Upon parasite recurrences detected by qPCR and confirmed by microscopy, a complete 7-day artesunate regimen was administered again and the initial sample collections repeated (i.e., sampling at 0h, 1h, 2h,…). Participants continued their follow-up until completion of the study at day 90.

From each blood sample, 50 µl were transferred into Trizol reagent for RNA preservation and immediately stored in liquid Nitrogen on-site. Samples were transferred to the Institut Pasteur of Cambodia in Phnom Penh and stored at −80°C until shipment to University of Maryland School of Medicine on dry ice. For whole-genome sequencing of parasites, blood collected before the first dose of artesunate (upon enrollment and in case of relapses) was passed on a column of cellulose to deplete white blood cells (WBC) and stored at −80°C until shipment to the University of Maryland on dry ice.

Thick and thin blood smears were made on microscopy slides from each blood sample collected and stained by Giemsa (3%, 45 min). Samples were considered negative if no parasite was detected after counting 500 WBCs on the thick films. Parasitemia was determined by counting the number of parasites for 500 WBCs and normalized using the estimate of 8,000 WBCs per microliter of blood. Parasite counts were used to determine the time to reduce by 99% the initial parasitemia (PCT99) as well as the slope half-life following regression models of the log-transformed parasite counts using the WWARN PCE online tool (www.wwarn.org/PCE).

### DNA extraction and whole genome sequencing

We extracted parasite DNA from leukocyte-depleted blood samples using the DNeasy blood and tissue kit (Qiagen) kit and prepared whole genome sequencing libraries using the NEBNext® Ultra™ II FS DNA Library Prep Kit for Illumina sequencing (NEB) according to the manufacturers’ instructions. We sequenced each library on an Illumina NovaSeq 6000 to generate 23-50 million paired-end reads of 100 bp per sample and mapped the reads from each sample onto the *P. vivax* P01 reference genome sequence [20] using *Bowtie2* [21].

### Complexity of Infection

To determine whether an infection was monoclonal, we assessed the level of nucleotide diversity within each sample. For this specific analysis, we remapped all reads to the *P. vivax* P01 v1 reference genome [20] for which the MalariaGEN Pv4 provides annotation of complex regions that can be masked to reduce mismapping artefacts [23]. We then used *GATK GenotypeGVCFs* [22] to call single nucleotide variants, only considering nucleotide positions that had a maximum of two alleles across all samples, no more than 20% missing information, and were located in the nuclear genome and outside of *P. vivax* multi-gene families. Finally, we estimated F_ws_ using *moimix* [23] and considered samples with F_ws_ >= 0.95 as monoclonal and those with F_ws_ < 0.95 as polyclonal.

To confirm these estimates of polyclonality and to evaluate the number of clones in polyclonal infections, as well as the proportion of the dominant clone (if any), we also examined the distribution of reference allele frequencies (RAF) within each sample [24]. Briefly, we used *samtools* [25] *mpileup* and custom scripts to determine, for each sample, the proportion of reference allele at each variable nucleotide position (excluding positions within multi-gene families). Samples for which most positions had a RAF close to 0 or 1 (i.e., all reads carrying the reference allele or all reads carrying the alternative allele) were considered monoclonal, while those with many positions displaying intermediate RAFs were considered polyclonal and the mode(s) of the distribution used to determine the relative proportions of the different clones [24].

### Genotype determination

To examine the relationships among different *P. vivax* clones, we used *samtool mpileup* and custom scripts to determine each parasite’s genotype at each variable position of the genome. For this analysis, we only considered variable positions that were sequenced at 20X or greater coverage in a given sample. Polyclonal samples were excluded, as well as one sample with >90% missing genotypes. Overall, we were able to analyze 33,686 variable nucleotide positions in 17 samples for this analysis. We then used *MEGA11* [26] to reconstruct a Neighbor-Joining tree based on the number of nucleotide differences between each pair of samples.

### Protein sequence analysis

We used *samtools* [25] *faidx* and *bcftools* [27] *consensus* to generate the DNA sequence from each sample (using genotype files from *GATK GenotypeGVCFs*) for ten *P. vivax* protein-coding genes (EPS15, UBP1, PATPL1, Coronin, K13, MRP2, PIP3, Ap2Mu, VP3 and falcilysin). We then used *TransDecoder* [28] to determine the corresponding amino acid sequence and *Mesquite* [29] to examine, separately for each gene, variations among isolates in amino acid sequences.

### RNA extraction and RNA-seq

We extracted RNA from blood samples collected in Trizol immediately upon collection using phenol-chloroform extraction. After rRNA depletion and polyA selection (NEB), we prepared RNA-seq libraries using the NEBNext Ultra II Directional RNA Library Prep Kit (NEB) and sequenced all libraries on an Illumina NovaSeq 6000 to generate ∼30-414 million paired-end reads of 75 bp per sample. We aligned all reads using *hisat2* [30] to a fasta file containing the *P. vivax* P01 and human hg38 genomes i) using default parameters and ii) using a shorter maximal intron length (--max-intronlen 5000). Reads mapping uniquely to the hg38 genome were selected from the BAM files generated with the default parameters. Reads mapping uniquely to the *P. vivax* genome were selected from the BAM files generated with a maximum intron length of 5,000 bp. PCR duplicates were removed from all files using custom scripts. We then calculated the number of reads mapped to each gene using annotations downloaded from PlasmoDB (*P. vivax* genes) and NCBI (human genes) and the *subread featureCounts* (v1.6.4) [31].

Read counts per gene were normalized into counts per million (cpm), separately for human and *P. vivax* genes. To filter out lowly expressed genes, only genes that were expressed at least at 10 cpm in > 50% of the samples were retained for further analyses (9,864 human and 4,923 *P. vivax* genes, respectively). Statistical assessment of differential expression was conducted, separately for the human and *P. vivax* genes, in *edgeR* (v 3.32.1) [32] using a quasi-likelihood negative binomial generalized model. All results were corrected for multiple testing using FDR [33].

We also estimated in each sample the proportion of each developmental stage using CIBERSORTx [34] and a custom signature matrix derived from orthologous *Plasmodium berghei* genes [35].

## Results

### Characteristics of the patients and infections treated with artesunate

We analyzed data from 87 vivax malaria patients enrolled in the Kampong Speu province, Western Cambodia, between November 2021 and December 2022. During the 90-day follow-up, 12 participants experienced one relapse, 11 had two relapses, 12 had three relapses and one had four relapses. Baseline characteristics of the participants and infections are shown in **Table 1**. Altogether, a total of 161 *P. vivax* mono-infections, confirmed by qPCR, were treated with a supervised 7-day course of artesunate and are analyzed here. In Cambodia, malaria transmission essentially occurs in forested areas and therefore predominantly affects young adult males engaging in forest activities. Accordingly, most participants were males (93%) with a median age of 20 years old. Neither the sex nor the age of the patient was significantly associated with the numbers of relapses observed within the 90-day monitoring period (respectively, p=0.672 and p=0.9034) (**Table 1**).

**Table 1.**
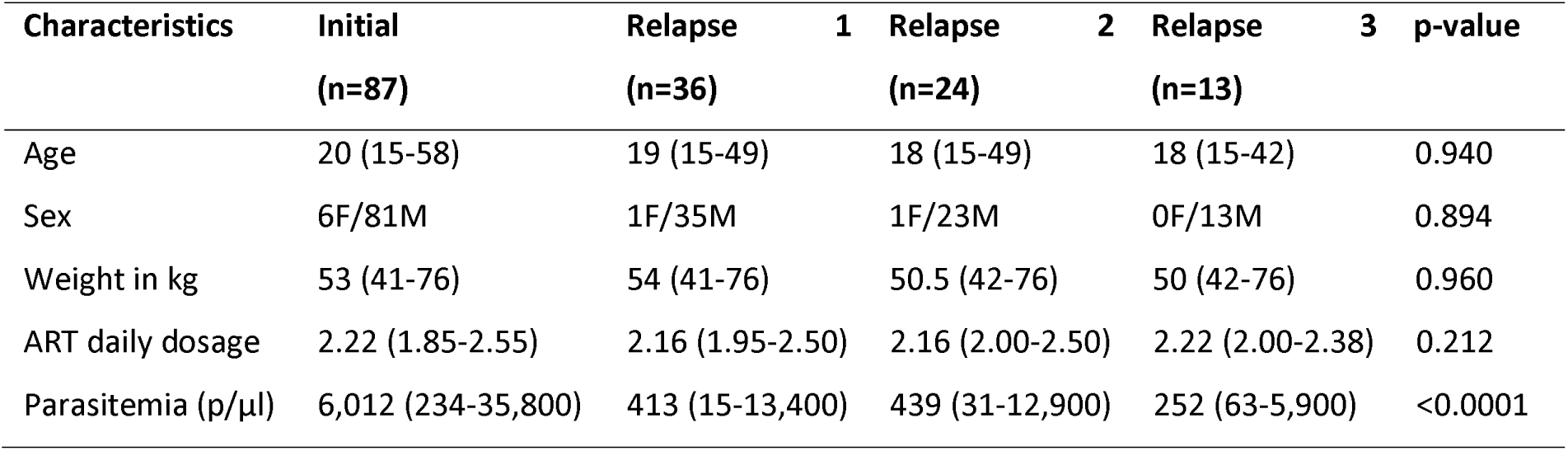
Participant characteristics and clinical information at the times of collection. The table displays the median values for each parameter with the range displayed in brackets. Data for the single relapse 4 infection is not presented.

Each participant received 1.85 to 2.55 mg of artesunate per kg per day for seven days (mean ± SD: 2.21 ± 0.15) depending on his/her weight (**Table 1**). Neither the patient’s weight, nor the actual dose of artesunate taken were significantly associated with the numbers of relapses observed within the 90-day monitoring period (Kruskal-Wallis, p=0.973 and p=0.442, respectively, **Supplemental Figure 1**).

Parasitemia upon initial enrollment ranged from 234 to 35,800 parasites/µl of blood (mean ± SD: 8,531 ± 7691). Since the relapses were treated as soon as parasites were detected by microscopy, their parasitemia were significantly lower than those of the infections at enrollment, when the participants seek malaria treatment (Kruskal-Wallis, p<0.0001, **Table 1**, **Supplemental Figure 1**). Parasitemia of the first, second or third relapses were not significantly different from each other (Kruskal-Wallis, p=0.746, **Table 1**).

### Determination of *in vivo* parasite clearance after artesunate treatment

All infections were successfully cleared by the artesunate treatment and the patients were deemed free of parasites by day 3 and 6 based on microscopy and PCR, respectively (**Table 2**). The effectiveness of artemisinin at treating *P. vivax* blood-stage infections was further demonstrated by the observation that, out of 46 patients included in the overall study and treated with a fourteen-day regimen of 0.5 mg/kg/day of primaquine after the completion of the artesunate treatment, only two patients showed reoccurrence of *P. vivax* parasites within the 90 day monitoring period (and those might derive from relapsing hypnozoites incompletely cleared by primaquine due to host genotypes)[36]. The times to achieve parasite clearance were significantly longer for initial infections compared to relapses (p=0.0005 using microscopy and p=0.0031 using qPCR), probably due to the higher parasitemia at enrollment (see also below).

**Table 2.**
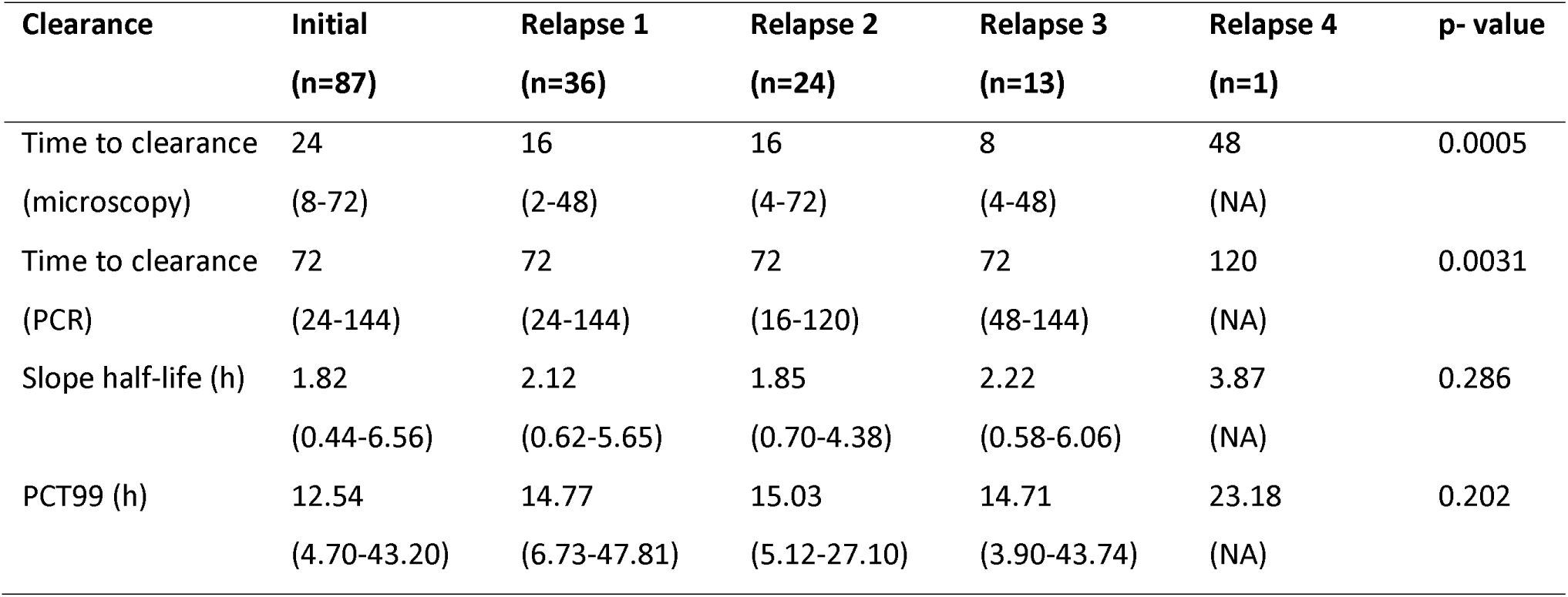
Clearance times and rates for the initial and relapse infections. The table displays the median values for each parameter with the range indicated in brackets.

We quantified parasite clearance rate by measuring, for each infection, i) the PCT99 (i.e., time to clear 99% of the parasites) and ii) the slope half-life (which takes into account the initial lag phase prior to exponential decline in parasite count after treatment [37]). Out of the 161 infections, PCT99 and slope half-life could be determined for 158 of them. As expected, PCT99 and the slope half-life were highly correlated with each other (Spearman, r=0.9672, P<0.0001, **Supplemental Figure 2**).

Interestingly, the distribution of slope half-lives measured for the 158 *P. vivax* infections showed a long tail, possibly indicative of heterogeneity in the parasites’ response to artesunate (**Figure 1A**). Indeed, expectation-maximization model-based clustering indicated that the data was best explained by a mixture of two gaussian distributions: one, accounting for 2/3 of the infections, with a mean of 1.602 and a second one with a mean of 3.298, **Supplemental Information**).

**Figure 1.**
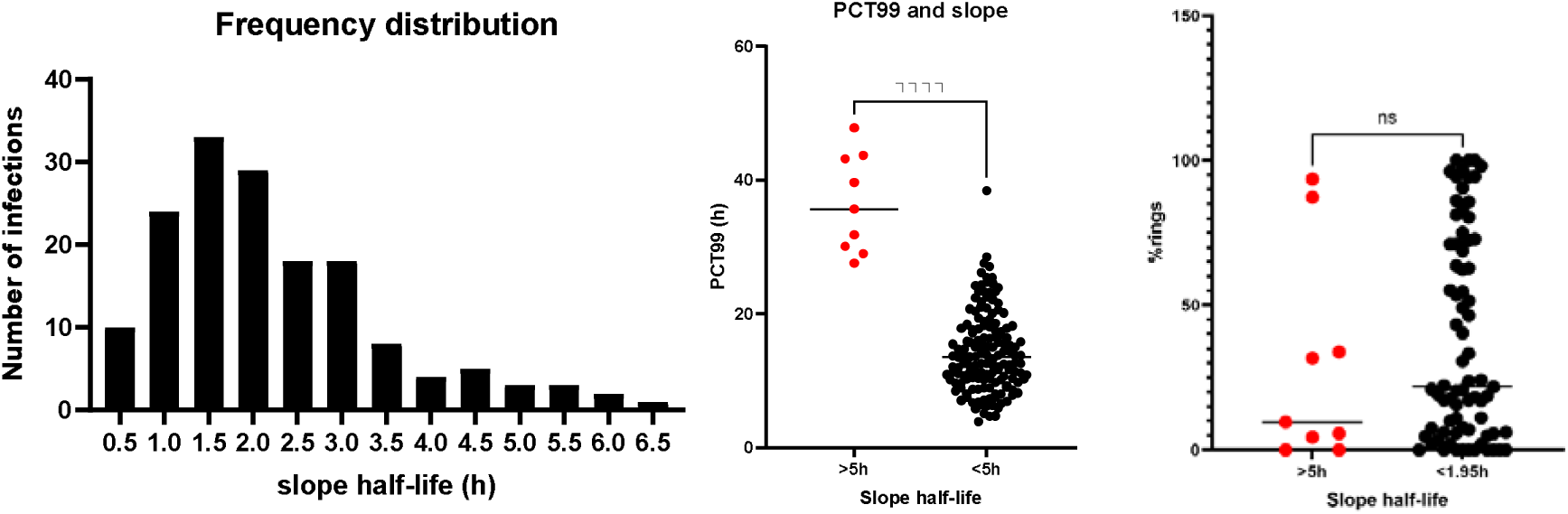
*In vivo* parasite clearance time after artesunate treatment. **(A)** Number of infections (y-axis) with a given slope half-life (x-axis, in hours). **(B)** PCT99 (y-axis) of each infection, shown separately for infections with a slope-half life of less and more than 5h (the WHO criteria for artemisinin resistance for *P. falciparum*). **(C)** Proportion of ring stage parasites in each infection determined by microscopy (y-axis, in %) according to the slope half-life after artesunate treatment.

Out of the 158 infections, nine (5.6%) showed a slope-half-life greater than 5h, the threshold defined in *P. falciparum* as indicative of *in vivo* resistance to artemisinin [12]. All nine infections with a prolonged slope half-life were also among the longest to clear, with a mean PCT99 of 36.5h, significantly longer than infections with slope half-life of less than 5h (mean of 14.3h, Mann Whitney p<0.0001, **Figure 1B**). Note also that one infection, that did not quite meet the resistance criteria with a slope half-life of 4.7h, displayed a long PCT99 (38.5h, **Figure 1B**).

### Parasite clearance times are not associated with characteristics of the infection or the host

There was no significant difference in the average PCT99 (Kruskal-Wallis, p=0.2070) and slope half-life (Kruskal-Wallis, p=0.3594) between infections at inclusion or upon first, second or third relapse (**Table 2, Supplemental Figure 3**). Similarly, there was no correlation between the slope half-lives and either i) parasitemia before the first dose of artesunate (Spearman, p= 0.2273), ii) patients’ age (p=0.3703), iii) patients’ weight (p=0.2621) or iv) the actual daily dosage of artesunate administered (p=0.8408) (**Supplemental Figure 4)**. There was also no difference in average parasitemia (Mann-Whitney, p=0.5149), patients’ age (p=0.1097), weight (p=0.4362) or daily dose of artesunate (p=0.3628) between infections with a slope half-life of more than 5h and those with a slope half-life below the median (<1.95h) (**Supplemental Figure 4**). Finally, there was no significant difference in the proportion of infections with a slope half-life of more than 5h between initial infections (5/89, 5.8%) and relapsing infections (4/67, 5.6%, Fisher p>0.999).

To determine whether host factors could be responsible for the delayed clearance phenotype (e.g., host immunity or host polymorphisms impacting artesunate metabolism or availability), we analyzed data from patients experiencing at least one relapse in addition to the initial infection. We observed no significant association between the slope half-lives and the patient ID (Fisher test, p=0.136). We also examined six patients that had multiple infections including at least one infection with a delayed clearance and all six patients displayed both slow and normal clearing infections (**Table 3**). Overall, these analyses suggest that the slow clearance of *P. vivax* parasites after artesunate treatment was not entirely driven by host factors but likely involved endogenous and/or environmental factors.

**Table 3.** Slope half-life of all *P. vivax* episodes for the subset of patients with multiple infections including at least one infection with a delayed clearance (indicated in red). Each line corresponds to a different patient.

| Patient ID | Initial | R1 | R2 | R3 | R4 |
| --- | --- | --- | --- | --- | --- |
| 1 | 0.48 | 2.98 | 1.34 | 5.16 | 3.87 |
| 2 | 2.28 | 1.04 | 0.70 | 6.06 |  |
| 3 | 1.00 | 5.65 | 3.18 | 3.22 |  |
| 4 | 5.42 | 5.44 | 3.50 | 3.11 |  |
| 25 | 5.09 | 1.76 |  |  |  |
| 26 | 6.56 | 3.54 |  |  |  |

Finally, we tested whether the stage composition of the infection before artesunate treatment influenced the clearance rate. We observed no correlation between the stage composition (summarized as the percentage of ring-stage parasites determined by microscopy) and the slope half-life (Spearman, p=0.8637). The percentage of ring stages in infections with a slope half-life greater than was also not significantly different than those with a slope half-life below the median (<1.95h) (Mann Whitney, p= 0.4134, **Figure 1C**).

### RNA sequencing of *P. vivax* parasites from human blood samples

We extracted and sequenced RNA from 32 *P. vivax* infections (23 infections at enrollment and 9 relapses), derived from 28 patients (**Supplemental Table 1A**). Eight of those infections had a clearance half-life greater than 5h and were considered “resistant”, following the WHO definition. A further eight infections had a clearance half-life between 3.5 and 5h and were considered “slow-clearing”. Finally, 15 infections had a clearance half-life lower than 1.95h (the median clearance time of the entire cohort) and were considered “fast-clearing”.

From each infection, we analyzed blood collected before treatment with artesunate, and 1, 2 and 4h after treatment, for a total of 111 blood samples (RNA-sequencing was not successful for every infection at every time point, **Supplemental Table 1A**).

We obtained between 30,875,718 and 417,614,734 read pairs per sample and between 4,956 and 127,094,184 (0.004% - 56.18%) reads mapped to *P. vivax* genome, while 1,741,494 to 264,706,126 (2.88% - 92.03%) reads mapped to the human genome.

### Slow- and fast clearing parasites have similar gene expression profiles before treatment

We first examined the gene expression profiles of all parasites before artesunate treatment. We used gene expression deconvolution to estimate, from each infection, the relative proportion of the different developmental stages [35]. Consistent with the microscopy analysis, we observed no statistically significant difference in the proportion of ring stage parasites in slow-vs. fast clearing infection before treatment (p = 0.54**, Supplemental Figure 5**).

Next, we used principal component analysis to examine similarities among parasite gene expression profiles. Overall, we failed to observe any obvious clustering of the parasites with similar clearance time before treatment (**Supplemental Figure 6A**) and neither PC1 nor PC2 was significantly correlated with the slope half-life (p = 0.18 and p = 0.44, respectively, **Supplemental Figure 6B/C**). We then tested whether specific parasite genes statistically differed in expression between fast- and slow-clearing infections, accounting for differences in stage composition. After correcting for multiple testing using false discovery rate [33], no genes remained significantly differentially expressed (FDR=0.1, **Supplemental Table 2**), indicating that slow- and fast-clearing parasites had similar gene expression profiles before artesunate treatment.

### Effect of artesunate on the gene expression of fast-clearing *P. vivax* parasites

We evaluated whether the proportion of each developmental stage present in the blood changed upon treatment using estimates from gene expression deconvolution [35]. In fast-clearing parasites, we observed no major differences in stage composition between 1h and 2h after treatment, with only a minor increase in the proportion of rings (from 24 to 26% on average, p=0.31) and a minor reduction in the proportion of trophozoites (from 37 to 31%, p=0.025) (**Supplemental Figure 7),** consistent with the stage-specific responses to ARTs described in *P. falciparum* [38].

To investigate the response of fast-clearing *P. vivax* parasites to artesunate, we analyzed the dynamics of gene expression changes by comparing the transcriptomes of parasites collected from the same infection at two consecutive time points (i.e., pre-treatment vs. 1h after artesunate treatment, 1h vs. 2h after treatment,…). Most changes in gene expression among fast-clearing *P. vivax* parasites occurred in the first two hours following treatment: 178 *P. vivax* genes displayed significant changes in gene expression between the pre-treatment sample and 1h after artesunate administration while 783 genes were differentially expressed genes between samples collected between 1h and 2h after treatment (FDR<0.1, **Figure 3A, Supplemental Table 3**). After 2h post-treatment, the parasite gene expression remained mostly unchanged, with 0 differentially expressed genes between 2h and 4h post treatment (**Figure 3A**).

**Figure 3:**
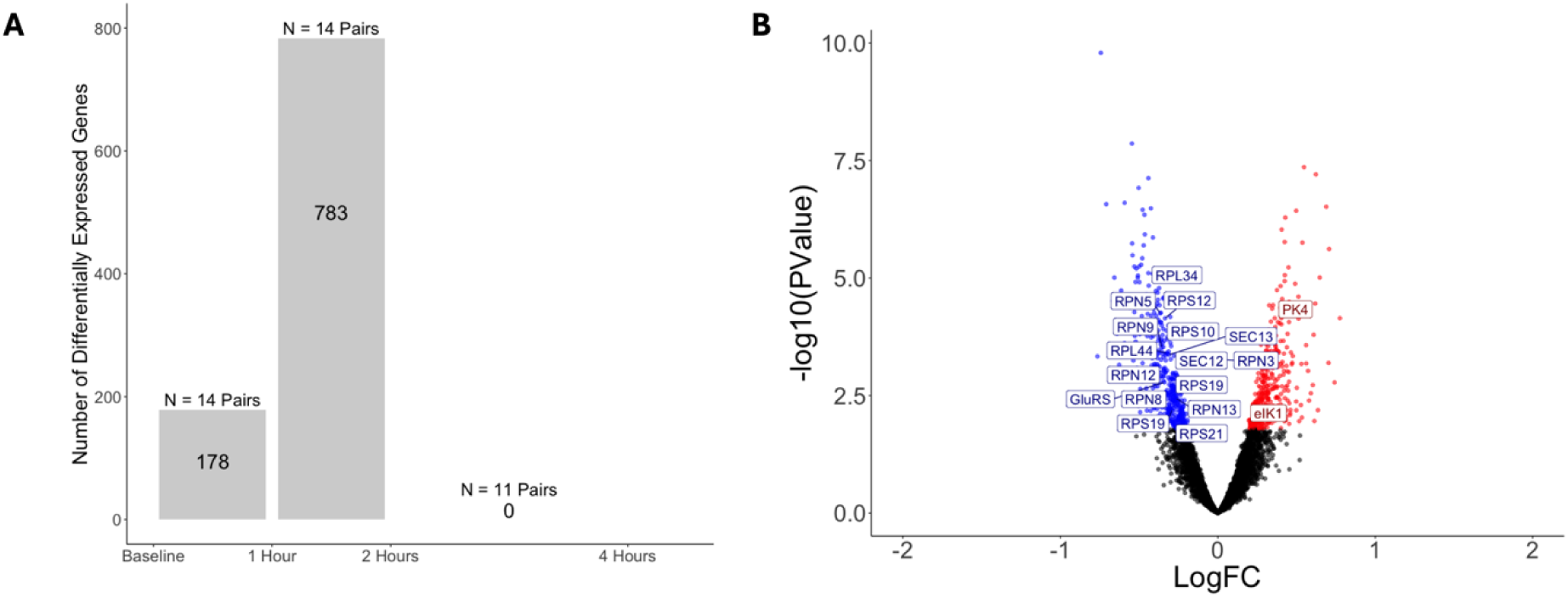
Summary of the changes in gene expression in fast-clearing parasites after artesunate treatment. **(A)** Each bar represents the number of differentially expressed *P. vivax* genes between the two time points (FDR=0.1). The numbers above each bar indicate the number of paired samples included in the analysis. **(B)** Volcano plot showing differentially expressed *P. vivax* genes between 1h and 2h post-artesunate treatment. Blue points indicate *P. vivax* genes that were significantly less expressed at 2h than 1h post-treatment, red points indicate genes that were significantly more expressed at 2h post-treatment. Blue gene labels highlight decreased expression of some proteasome and translation-related genes.

In both comparisons, before vs. 1h post-artesunate treatment and 1h vs. 2h post treatment, many differentially expressed genes were part of the proteasome complex pathway: three protease/proteasome subunits showed lower expression 1h after treatment than at baseline, while ten subunits had lower expression 2h post-treatment than 1h post-treatment (**Supplemental Table 3**). In addition, one and five ubiquitin-related genes were differentially expressed between these time points, respectively. We also observed a strong signal consistent with an overall downregulation of translation: six ribosomal proteins had lower expression 1h post-treatment than before treatment (while no ribosomal proteins increased in expression) and this effect was magnified in the 1h vs. 2h comparison where 20 ribosomal proteins showed decreased expression (for only three with increased expression). These patterns mirror the effects of artemisinin described for *P. falciparum*, where prior work identified decreased expression of proteasomal [12] and ribosomal genes [12, 38] upon artesunate exposure.

Interestingly, the expression of many genes involved in antioxidant defense [39] also decreased upon artesunate treatment, including superoxide dismutase (PVP01_1126000), 1-cys peroxiredoxin (PVP01_0118000), and glutathione synthetase (PVP01_1021800).

### Delayed gene expression response of slow-clearing *P. vivax* parasites upon treatment

We then analyzed the gene expression of the slow-clearing and resistant parasites (half-life > 3.5h, referred to jointly as slow-clearing for the rest of the analyses). The gene expression profiles of these parasites did not reveal any change in stage composition between samples collected before artesunate treatment and 1h post treatment but showed a significant decrease in the proportion of trophozoites between 1h and 2h post treatment (from 53 to 49%, p=0.038) and this decrease continued until the 4h time point (with a final proportion of 31%, p=2.7×10^-5^) (**Supplemental Figure 8**). The proportion of male gametocytes increased between 1h and 2h post treatment (from 3% to 4%, p = 0.029) and the proportion of female gametocytes increased between 2h and 4h post treatment (from 12% to 27%, p = 0.002) (**Supplemental Figure 8**).

We then examined the dynamics of the gene expression changes of slow-clearing *P. vivax* parasites upon artesunate treatment, again comparing the transcriptomes of parasites collected from the same infections at two consecutive time points. In contrast to fast-clearing parasites, we identified only one gene that changed expression between the pre-treatment samples and 1h post-treatment. Changes in gene expression only became detectable 2h post-artesunate, when we identified 1,703 differentially expressed genes (with 476 of them also differentially in the fast-clearing parasites), and peaked between 2h and 4h post-artesunate, with 2,445 differentially expressed genes, of which all were unique to slow-clearing parasites **(Figure 4A, Supplemental Table 3)**. Overall, these results indicated both a shared but delayed gene expression response to artesunate treatment of the slow-clearing parasites, as well as a unique transcriptional signature, specific to these slow-clearing parasites.

**Figure 4:**
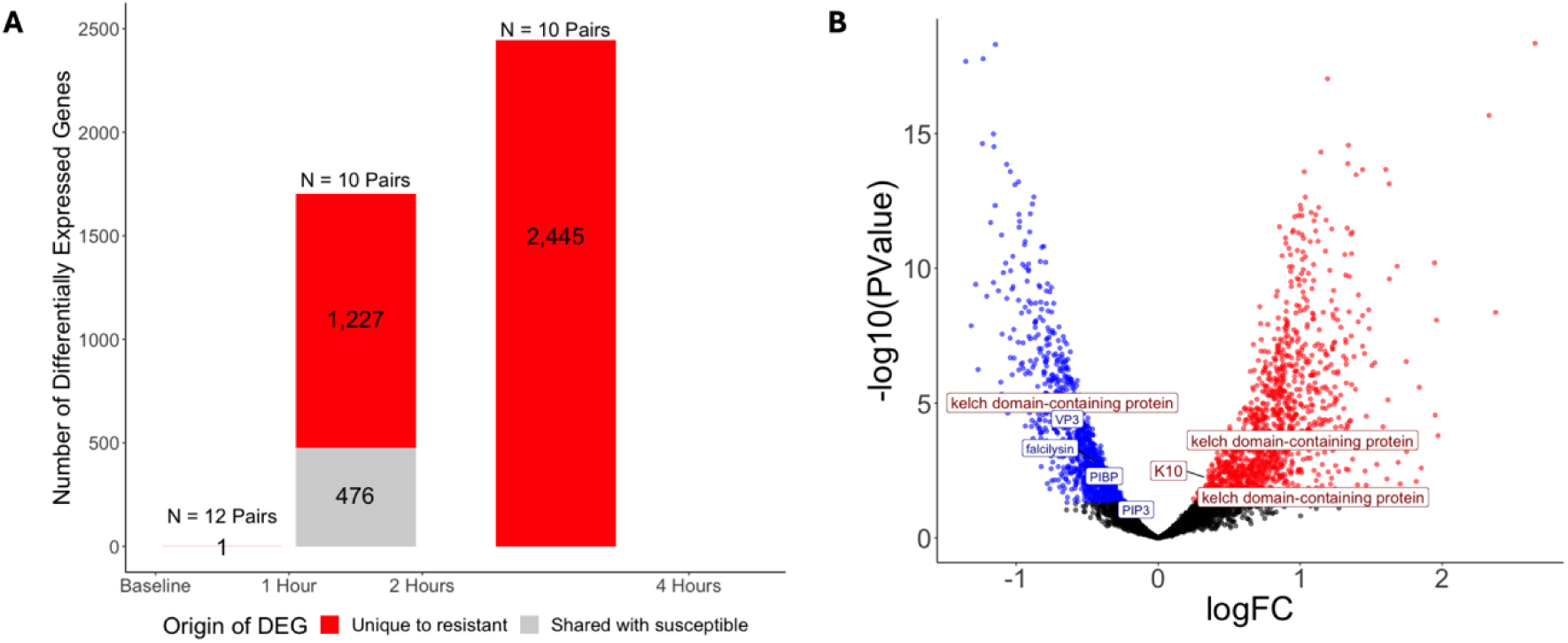
Summary of differential gene expression in slow-clearing parasites. **(A)** Each bar represents the number of differentially expressed *P. vivax* genes between the two time points (FDR=0.1). Red segments represent genes that are uniquely differentially expressed in slow-clearing parasites while the grey segments showed genes that are differentially expressed in both slow- and fast-clearing parasites. Numbers above each bar indicate the number of sample pairs included in each comparison. **(B)** Volcano plot showing differential gene expression between 2h and 4h post-artesunate treatment. Blue points indicate genes that are significantly decreased in expression at 4h post-treatment, red points indicate genes that are significantly increased in expression at 4h post-treatment. Blue labels highlight genes involved in the hemoglobin digestion pathway, red labels highlight kelch-domain containing genes.

Out of the 4,800 differentially expressed genes in slow-clearing parasites, 476 (1%) were also differentially expressed in fast-clearing parasites (and those represented 97% of all genes differentially expressed in fast-clearing). These include many genes related to the proteasome, ribosome and translation and protein processing **(Supplemental Table 4)**.

Many of the genes differentially expressed uniquely in slow-clearing parasites were also associated with these pathways, suggesting that the slowing down of the proteosome and translational machinery observed in fast-clearing parasites upon artesunate treatment was magnified in slow-clearing parasites. Thus, 29 genes differentially expressed in slow-clearing parasites were associated with proteosome/ubiquitination pathways (vs. 15 in fast-clearing parasites) while 64 were involved in the regulation of translation (vs. 20) (**Supplemental Table 4**).

In addition to genes in these shared pathways, several differentially expressed genes pointed to unique metabolic responses occurring in the slow-clearing parasites. In particular, we noted that several genes essential for hemoglobin digestion were uniquely downregulated in slow-clearing parasites (e.g., **Figure 4B**). Those included both genes involved in hemoglobin endocytosis (e.g., VSP45 and phosphatidylinositol 3-kinase) and genes involved in hemoglobin catabolism (e.g., falcilysin and vivapain 3). This observation is particularly interesting since artesunate is activated by the heme produced during hemoglobin digestion, and parasites with slower hemoglobin digestion could therefore be subjected to less activated drug [11, 40, 41]. While several kelch domain-containing proteins (PVP01_1304500, PVP01_0607800, PVP01_0926500, PVP01_0926400, PVP01_0932400, PVP01_1432200) were differentially expressed in slow-clearing parasites, we did not observe differential expression of the ortholog of Kelch13 that has been associated with artemisinin resistance in *P. falciparum* [42] (PVP01_1211100, qvalue=0.8).

Finally, we detected increased expression of AP2-G (PVP01_1440800) and AP2-G5 (PVP01_0940100) 4h after treatment. AP2-G is the master regulator of gametocyte development in *Plasmodium* species while AP2-G5 is a transcription factor responsible for the early development of gametocytes [43]. These observations could be consistent with an increase of sexual commitment upon artesunate treatment and/or an increased release of gametocyte from the bone marrow (that may explain the modest increase in gametocyte proportion detected by gene expression deconvolution, **Supplemental Figure 8**). Similar patterns have been described in *P. falciparum* both *in vitro* [44] and in human patients [45], where artesunate treatment leads to an increase in the proportion of sexual stages. This observation, that will need to be validated, could have worrying implications as it suggests that artesunate treatment could increase parasite transmission (especially since *P. vivax* has a more rapid gametocytogenesis than *P. falciparum*).

### Effect of artesunate treatment on human gene expression

Finally, we analyzed the effect of artesunate treatment on the host gene expression profiles and tested whether the differences between fast- and slow-clearing parasites may be the result of differences in the human immune response. First, we tested for differences in host gene expression between the fast- and slow clearing infections and identified no genes differentially expressed before treatment (FDR=0.1, **Supplemental Figure 9A**). Second, we examined whether the expression of specific human genes changed upon artesunate treatment. We only identified 62 differentially expressed human genes one hour after treatment in the fast-clearing infections and none in the slow-clearing infections (**Supplemental Figure 9B/C**). Those include genes related to the innate immune response (e.g., CD300C) and regulation of the immune system (LILRB isoforms), that were increased in expression after treatment, and genes related to lymphocyte development and activation (e.g IL7R) that were decreased in expression. Overall, these gene expression differences, especially compared to the much stronger signal observed for parasite genes, seem relatively minor and possibly only result from a change in inflammation in response to extensive parasite death caused by the artesunate treatment.

### Whole genome sequencing of *P. vivax* parasites and association with clearance time

We extracted DNA and sequenced the genomes of *P. vivax* parasites from 30 of the infections analyzed by RNA-seq after artesunate treatment (21 infections at enrollment and 9 upon relapses, **Supplemental Table 1B**). On average, ∼40% of the reads generated from each sample mapped uniquely to the *P. vivax* reference genome (ranging from 0.07% to 79.23%), providing high coverage genome data (>30X) for 28 infections that were further analyzed (**Supplemental Table 1B**).

We first used the data generated to examine variations in protein-coding sequences at genes possibly involved in the slow-clearance upon artesunate treatment based on i) results from resistance studies conducted in *P. falciparum* (e.g., Kelch13) and ii) candidates based on our gene expression findings (e.g., falcilysin, vivapain). Due to the relatively small number of isolates analyzed here, we focused on only 10 candidate genes and exclusively examined coding polymorphisms. Overall, there were no obvious missense or non-synonymous polymorphisms differentiating fast- from slow-clearing isolates (p>0.07, **Supplemental Table 4**).

We also leveraged the whole genome sequencing data to determine the complexity of each infection. 15 out of the 28 infections sequenced at sufficient coverage were deemed monoclonal (Fws>0.95). For the 15 monoclonal infections, and 2 polyclonal infections in which one clone clearly dominated and accounted for more than 60% of the parasites, we identified 33,686 single nucleotide variants and used these polymorphisms to examine genetic relationships among these different isolates (**Figure 5**). Most isolates were equally distant from all other parasites, as previously described from population sampling [46, 47]. In particular, slow-clearing parasites did not appear to cluster closely together as one would expect if they all shared a recent beneficial mutation enabling them to overcome artesunate treatment. Performing this analysis per chromosome (which would reduce the diluting effect of recombination) did not affect qualitatively these results. Similarly to what has been previously described [48–51], we observed that relapsing parasites could be unrelated, meiotic siblings (e.g., infection 39 on Figure 5) or identical. Interestingly, the parasites that reoccurred in patient 12 44 days after the initial infection, fell in this latter category and were genetically identical to those at enrollment. However, while the initial infection cleared rapidly (with a half-life of 1.76), the relapsing parasites were eliminated much slower (with a half-life of 4.6h). This singular observation suggested that the slow clearance of *P. vivax* parasites might not be genetically regulated (e.g., by a resistant mutation) but instead may be determined by environmental or epigenetic factors.

**Figure 5:**
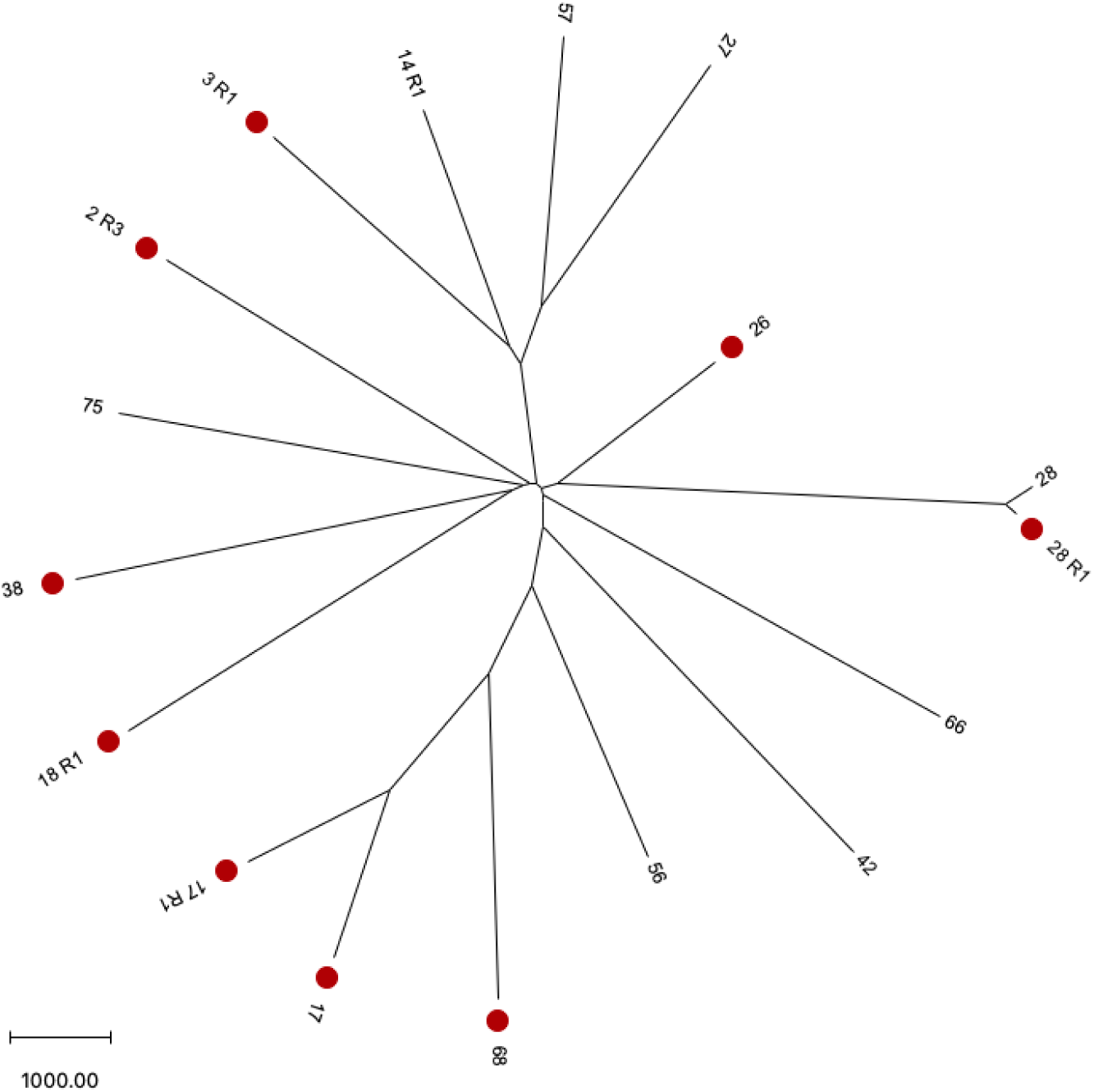
Relationships among *P. vivax* isolates included in this study. The figure shows a neighbor-joining tree based on the number of nucleotide differences between each pair of samples (using 33,686 variable positions determined from whole genome sequencing). Parasites from slow-clearing infections are labelled in red, while parasites from fast-clearing infections are not highlighted. Note that the same parasite genotype is present in both one fast- and one slow-clearing infection (from patient 12 at D0 and a relapse at D44, respectively).

## Conclusion

We characterized, for the first-time, the response of *P. vivax* parasites to artesunate using Cambodian vivax malaria patients receiving supervised artesunate treatment. All patients were cleared of parasites by day 3 of artesunate monotherapy treatment and we did not observe any indication of treatment failure. Nonetheless, we observed a wide range of clearance times among infections and nine infections displayed a clearance half-life greater than 5h, the threshold used by the WHO for artemisinin resistance in *P. falciparum.* This first report of delayed clearance upon artesunate treatment in *P. vivax,* while not associated with treatment failure, is worrying as this slow-clearance may enable parasites to outlast artesunate treatment (especially in cases of incomplete treatment adherence) and, potentially, to develop resistance to partner drugs used in ACT. Gene expression analyses suggest that reduced hemoglobin digestion in slow-clearing parasites may explain this phenotype, as lesser hemoglobin digestion could impair efficacious activation of artesunate. In addition, genetic analyses indicate that slow-clearing parasites are not genetically closely related (as one would expect if these parasites shared a recently acquired mutation enabling this slow-clearance) and do not carry obvious mutations in the coding sequences of key artesunate resistance genes (although larger studies will be necessary to confirm these results). Instead, we speculate that environmental factors, such as the host nutritional status, or epigenetic factors may regulate the growth rate of *P. vivax* parasites and, consequently, the efficacy or artesunate at clearing an infection.

## Acknowledgements

We thank all patients and health care workers involved in this study and the staff of the Malaria Research Unit at the Institut Pasteur in Cambodia and the staff of the National Center for Parasitology, Entomology and Malaria Control in Cambodia for their collaboration and sample collection. We thank Drs. Baird, Lin and Nosten for the suggestions and advice they provided throughout the study. We also thank S. Ott, H. Bowen, L. Sadzewicz, and L. Tallon in Maryland Genomics at the University of Maryland School of Medicine for their support with Illumina sequencing. This study was supported by an award from the NIH (R01AI146590). JP is supported by the NIH/NIAID (R01AI173171, R01AI175134 and R61AI187100) and by the Pasteur International Unit PvESMEE. The funders had no role in study design, data collection and analysis, decision to publish, or preparation of the manuscript.

## Data and code availability

All sequence data generated in this study have been deposited in the National Center for Biotechnology Information (NCBI) Sequence Read Archive under the BioProject ID XXXX. Custom scripts are available at https://github.com/tebbenk/PvARTr.

## Supplemental Figure legends

**Supplemental Figure 1.** Relapse characteristics. The figure shows that weight, age and actual artesunate daily dose are not associated with the relapses but that relapses have a significantly lower parasitemia.

**Supplemental Figure 2.** PCT99 and slope half-life are highly correlated with each other.

**Supplemental Figure 3.** Clearance times are similar at enrollment and for relapse infections.

**Supplemental Figure 4.** No patient or infection parameters are associated with clearance times. The figures show the lack of association between parasitemia, the patient age, the patient weight and the actual dose of artesunate administered and the clearance times.

**Supplemental Figure 5.** Comparison of the proportion of ring stage parasites estimated by gene expression deconvolution from fast- and slow-clearing infections.

**Supplemental Figure 6.** Principal component analysis of the parasite gene expression before artesunate treatment, with fast-clearing parasites indicated in black and slow-clearing parasites in grey **(A)**. Correlation between the two first principal components and the infection slope half-life (**B,C**).

**Supplemental Figure 7.** Differential gene expression and developmental stage composition between one-hour and two-hours post-artesunate treatment in fast-clearing parasites. (**A**) Volcano plot where each point represents one *P. vivax* gene plotted by its p-value (y-axis) and log fold-change (x-axis). Blue points represent genes that are significantly decreased in expression at one-hour post-treatment, red points represent genes that are significantly increased in expression at one-hour post-treatment. (**B-F**) Boxplots depicting the distribution of the relative proportions of rings (B), trophozoites (C), schizonts (D), female gametocytes (E) and male gametocytes (F) present in each sample at one- and two-hours post-treatment estimated by gene expression deconvolution. Each box displays the 25th percentile value, median, and 75th percentile value.

**Supplemental Figure 8.** Differential gene expression and developmental stage composition post-artesunate treatment in slow-clearing parasites. (A) Volcano plot of *P. vivax* genes between 1 h and 2 h post-treatment where each point represents one gene plotted by its p-value (y-axis) and log fold-change (x-axis). Blue points represent genes that are significantly decreased in expression at 2 h post-treatment. Red points represent genes that are significantly increased in expression at 2 h post-treatment. (**B-F**) Boxplots depicting the distribution of the relative proportions of rings (B), trophozoites (C), schizonts (D), male gametocytes (E) and female gametocytes (F) present in each sample at 1 h and 2 h post-treatment estimated by gene expression deconvolution. Each box displays the 25th percentile value, median, and 75th percentile value. (**G**) Volcano plot of P. vivax genes between 2 h and 4 h post-treatment. Blue points represent genes that are significantly decreased in expression at 4 h post-treatment. Red points represent genes that are significantly increased in expression at 4 h post-treatment. (**H-L**) Boxplots depicting the distribution of the relative proportions of rings (H), trophozoites (I), schizonts (J), male gametocytes (K) and female gametocytes (L) present in each sample at baseline and one-hour post-treatment estimated by gene expression deconvolution. Each box displays the 25th percentile value, median, and 75th percentile value.

**Supplemental Figure 9.** Human differential gene expression after treatment with artesunate. Volcano plots where each point represents one human gene plotted by its p-value (y-axis) and log fold-change (x-axis). (**A**) Human differential gene expression between infections with fast- and slow-clearing parasites before treatment. (**B**) Human differential gene expression of infections with fast-clearing parasites between baseline and one-hour post-treatment. (**C**) Human differential gene expression of infections with slow-clearing parasites between baseline and one-hour post-treatment. Blue points represent genes that are significantly decreased in expression at one-hour post-treatment. Red points represent genes that are significantly increased in expression at one-hour post-treatment.

**Supplemental Text.**
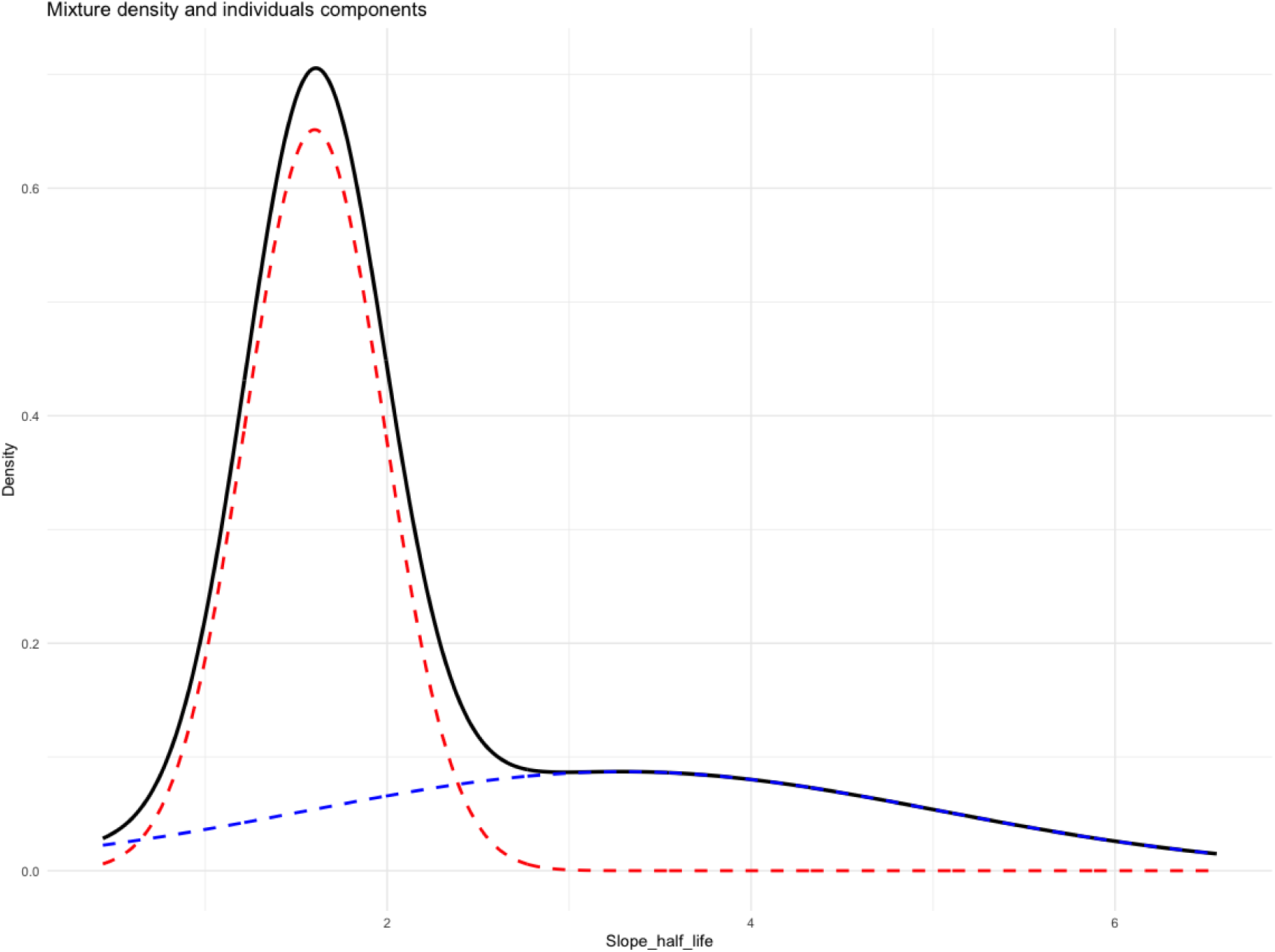
To estimate whether the distribution of slope half-life for all infections (Figure 1A) was best explained by a single gaussian distribution, we used an Expectation-Maximization algorithm initialized by hierarchical model-based agglomerative clustering. The optimal model (i.e., the optimal numbers of different normal distribution) was selected according to BIC using the R package *mclust* (R 4.3.3). The data best fitted a mixture model composed of two gaussian distributions: the first distribution accounting for 109 infections (with a mean of 1.602 h and a variance of 0.38, in red) and the second accounting for 49 infections (with a mean of 3.298 h and a variance of 1.737, in blue).

## References

1. Dondorp, A.M., et al., Artemisinin resistance in Plasmodium falciparum malaria. N Engl J Med, 2009. 361(5): p. 455–67.

2. Noedl, H., et al., Evidence of artemisinin-resistant malaria in western Cambodia. N Engl J Med, 2008. 359(24): p. 2619–20.

3. Ashley, E.A., et al., Spread of artemisinin resistance in Plasmodium falciparum malaria. N Engl J Med, 2014. 371(5): p. 411–23.

4. Rosenthal, P.J., et al., The emergence of artemisinin partial resistance in Africa: how do we respond? Lancet Infect Dis, 2024.

5. Uwimana, A., et al., Association of Plasmodium falciparum kelch13 R561H genotypes with delayed parasite clearance in Rwanda: an open-label, single-arm, multicentre, therapeutic efficacy study. Lancet Infect Dis, 2021. 21(8): p. 1120–1128.

6. Balikagala, B., et al., Evidence of Artemisinin-Resistant Malaria in Africa. N Engl J Med, 2021. 385(13): p. 1163–1171.

7. Agrawal, S., et al., Association of a Novel Mutation in the Plasmodium falciparum Chloroquine Resistance Transporter With Decreased Piperaquine Sensitivity. J Infect Dis, 2017. 216(4): p. 468–476.

8. Amaratunga, C., et al., Dihydroartemisinin-piperaquine resistance in Plasmodium falciparum malaria in Cambodia: a multisite prospective cohort study. Lancet Infect Dis, 2016. 16(3): p. 357–65.

9. Amato, R., et al., Genetic markers associated with dihydroartemisinin-piperaquine failure in Plasmodium falciparum malaria in Cambodia: a genotype-phenotype association study. Lancet Infect Dis, 2017. 17(2): p. 164–173.

10. Golenser, J., et al., Current perspectives on the mechanism of action of artemisinins. Int J Parasitol, 2006. 36(14): p. 1427–41.

11. Tilley, L., et al., Artemisinin Action and Resistance in Plasmodium falciparum. Trends Parasitol, 2016. 32(9): p. 682–696.

12. Zhu, L., et al., Artemisinin resistance in the malaria parasite, Plasmodium falciparum, originates from its initial transcriptional response. Commun Biol, 2022. 5(1): p. 274.

13. Bridgford, J.L., et al., Artemisinin kills malaria parasites by damaging proteins and inhibiting the proteasome. Nat Commun, 2018. 9(1): p. 3801.

14. Ward, K.E., D.A. Fidock, and J.L. Bridgford, Plasmodium falciparum resistance to artemisinin-based combination therapies. Curr Opin Microbiol, 2022. 69: p. 102193.

15. Meshnick, S.R., Artemisinin: mechanisms of action, resistance and toxicity. Int J Parasitol, 2002. 32(13): p. 1655–60.

16. World Health Organization, World malaria report 2023. 2023, World Health Organization: Geneva.

17. Popovici, J., et al., Recrudescence, Reinfection, or Relapse? A More Rigorous Framework to Assess Chloroquine Efficacy for Plasmodium vivax Malaria. J Infect Dis, 2019. 219(2): p. 315–322.

18. Ferreira, M.U., et al., Monitoring Plasmodium vivax resistance to antimalarials: Persisting challenges and future directions. Int J Parasitol Drugs Drug Resist, 2021. 15: p. 9–24.

19. Price, R.N., et al., Global extent of chloroquine-resistant Plasmodium vivax: a systematic review and meta-analysis. Lancet Infect Dis, 2014. 14(10): p. 982–91.

20. Auburn, S., et al., A new Plasmodium vivax reference sequence with improved assembly of the subtelomeres reveals an abundance of pir genes. Wellcome Open Res, 2016. 1: p. 4.

21. Langmead, B. and S.L. Salzberg, Fast gapped-read alignment with Bowtie 2. Nat Methods, 2012. 9(4): p. 357–9.

22. Van der Auwera GA. and O.C. BD., Genomics in the Cloud: Using Docker, GATK, and WDL in Terra. 1 ed. 2020: O’Reilly Media.

23. Lee, S. and M. Bahlo, moimix: an R package for assessing clonality in high-througput sequencing data. 2016.

24. Chan, E.R., et al., Whole Genome Sequencing of Field Isolates Provides Robust Characterization of Genetic Diversity in Plasmodium vivax, in PLoS Neglected Tropical Diseases. 2012.

25. Li, H., et al., The Sequence Alignment/Map format and SAMtools. Bioinformatics, 2009. 25(16): p. 2078–9.

26. Tamura, K., G. Stecher, and S. Kumar, MEGA11: Molecular Evolutionary Genetics Analysis Version 11. Mol Biol Evol, 2021. 38(7): p. 3022–3027.

27. Li, H., A statistical framework for SNP calling, mutation discovery, association mapping and population genetical parameter estimation from sequencing data. Bioinformatics, 2011. 27(21): p. 2987–93.

28. Haas, B.

29. Maddison, W.P. and D.R. Maddison, Mesquite: A modular system for evolutionary analysis. Version 3.81. 2023.

30. Kim, D., B. Langmead, and S.L. Salzberg, HISAT: a fast spliced aligner with low memory requirements. Nat Methods, 2015. 12(4): p. 357–60.

31. Liao, Y., G.K. Smyth, and W. Shi, featureCounts: an efficient general purpose program for assigning sequence reads to genomic features. Bioinformatics, 2014. 30(7): p. 923–30.

32. Robinson, M.D., D.J. McCarthy, and G.K. Smyth, edgeR: a Bioconductor package for differential expression analysis of digital gene expression data. Bioinformatics, 2010. 26(1): p. 139–40.

33. Benjamini, Y. and Y. Hochberg, Controlling the False Discovery Rate: A Practical and Powerful Approach to Multiple Testing. Journal of the Royal Statistical Society. Serie B, 1995. 57(1): p. 289–300.

34. Newman, A.M., et al., Determining cell type abundance and expression from bulk tissues with digital cytometry. Nat Biotechnol, 2019. 37(7): p. 773–782.

35. Tebben, K., A. Dia, and D. Serre, Determination of the Stage Composition of Plasmodium Infections from Bulk Gene Expression Data. mSystems, 2022. 7(4): p. e0025822.

36. Eng, V., et al., High versus low dose of 14 days treatment of primaquine in Plasmodium vivax infected patients in Cambodia: a randomised open-label efficacy study. medRxiv, 2025: p. 2025.01.01.25319862.

37. Flegg, J.A., et al., Standardizing the measurement of parasite clearance in falciparum malaria: the parasite clearance estimator. Malar J, 2011. 10: p. 339.

38. Shaw, P.J., et al., Plasmodium parasites mount an arrest response to dihydroartemisinin, as revealed by whole transcriptome shotgun sequencing (RNA-seq) and microarray study. BMC Genomics, 2015. 16: p. 830.

39. Bozdech, Z. and H. Ginsburg, Antioxidant defense in Plasmodium falciparum--data mining of the transcriptome. Malar J, 2004. 3: p. 23.

40. Rosenthal, M.R. and C.L. Ng, Plasmodium falciparum Artemisinin Resistance: The Effect of Heme, Protein Damage, and Parasite Cell Stress Response. ACS Infect Dis, 2020. 6(7): p. 1599–1614.

41. Ullah, I., et al., Artemisinin resistance mutations in Pfcoronin impede hemoglobin uptake. bioRxiv, 2024.

42. Ariey, F., et al., A molecular marker of artemisinin-resistant Plasmodium falciparum malaria. Nature, 2014. 505(7481): p. 50–5.

43. Shang, X., et al., A cascade of transcriptional repression determines sexual commitment and development in Plasmodium falciparum. Nucleic Acids Res, 2021. 49(16): p. 9264–9279.

44. Portugaliza, H.P., et al., Artemisinin exposure at the ring or trophozoite stage impacts Plasmodium falciparum sexual conversion differently. Elife, 2020. 9.

45. Portugaliza, H.P., et al., Plasmodium falciparum sexual conversion rates can be affected by artemisinin-based treatment in naturally infected malaria patients. EBioMedicine, 2022. 83: p. 104198.

46. Chan, E.R., et al., Whole genome sequencing of field isolates provides robust characterization of genetic diversity in Plasmodium vivax. PLoS Negl Trop Dis, 2012. 6(9): p. e1811.

47. Pearson, R.D., et al., Genomic analysis of local variation and recent evolution in Plasmodium vivax. Nat Genet, 2016. 48(8): p. 959–964.

48. Popovici, J., et al., Genomic Analyses Reveal the Common Occurrence and Complexity of Plasmodium vivax Relapses in Cambodia. mBio, 2018. 9(1).

49. Imwong, M., et al., Relapses of Plasmodium vivax infection usually result from activation of heterologous hypnozoites. J Infect Dis, 2007. 195(7): p. 927–33.

50. Kim, J.R., et al., Genotyping of Plasmodium vivax reveals both short and long latency relapse patterns in Kolkata. PLoS One, 2012. 7(7): p. e39645.

51. Noviyanti, R., et al., Hypnozoite depletion in successive Plasmodium vivax relapses. PLoS Negl Trop Dis, 2022. 16(7): p. e0010648.

